# Low-cost measurement of facemask efficacy for filtering expelled droplets during speech

**DOI:** 10.1101/2020.06.19.20132969

**Authors:** Emma P. Fischer, Martin C. Fischer, David Grass, Isaac Henrion, Warren S. Warren, Eric Westman

## Abstract

Mandates for mask use in public during the recent COVID-19 pandemic, worsened by global shortage of commercial supplies, have led to widespread use of homemade masks and mask alternatives. It is assumed that wearing such masks reduces the likelihood for an infected person to spread the disease, but many of these mask designs have not been tested in practice. We have applied a simple optical measurement method to evaluate the efficacy of masks to reduce the transmission of respiratory droplets during regular speech. We compare a variety of commonly available mask types and observe that some mask types approach the performance of standard surgical masks, while some mask alternatives, such as neck fleece or bandanas, offer very little protection. Our measurement setup is inexpensive and can be built and operated by non-experts, allowing for rapid evaluation of mask performance during speech, sneezing, or coughing.

## Introduction

The global spread of COVID-19 in early 2020 has significantly increased the demand for face masks around the world, while stimulating research about their efficacy. Here we adapt a recently demonstrated optical imaging approach [1, 2] to highlight stark differences in the effectiveness of different masks and mask alternatives to suppress the spread of respiratory droplets during regular speech.

In general, the term ‘face mask’ governs a wide range of protective equipment with the primary function of reducing the transmission of particles or droplets. The most common application in modern medicine is to provide protection *to* the wearer (e.g. first responders), but surgical face masks were originally introduced to protect surrounding persons *from* the wearer, such as protecting patients with open wounds against infectious agents from the surgical team [3], or the persons surrounding a tuberculosis patient from contracting the disease via airborne droplets [4]. This latter role has been embraced by multiple governments and regulatory agencies [5], since COVID-19 patients can be asymptomatic but contagious for many days [6]. The premise of protection from infected persons wearing a mask is simple: wearing a face mask will reduce the spread of respiratory droplets containing viruses. In fact, recent studies suggest that wearing face masks reduces the spread of COVID-19 on a population level, and consequently blunts the growth of the epidemic curve [7, 8]. Still, determining mask efficacy is a complex topic that is still an active field of research (see for example [9]), made even more complicated because the infection pathways for COVID-19 are not yet fully understood and are complicated by many factors such as the route of transmission, correct fit and usage of masks, and environmental variables. From a public policy perspective, shortages in supply for surgical face masks and N95 respirators, as well as concerns about their side effects and the discomfort of prolonged use [10], have led to public use of a variety of solutions which are generally less restrictive (such as homemade cotton masks or bandanas), but usually of unknown efficacy. While some textiles used for mask fabrication have been characterized [11], the performance of actual masks in a practical setting needs to be considered. The work we report here reflects a first attempt to improve evaluation; such results could be used to guide mask selection and purchase decisions.

A schematic and demonstration image are shown in **Fig. 1**. In brief, an operator wears a face mask and speaks into the direction of an expanded laser beam inside a dark enclosure. Droplets that propagate through the laser beam scatter light, which is recorded with a cell phone camera. A simple computer algorithm is used to count the droplets in the video. The required hardware for these measurements is commonly available; suitable lasers and optical components are accessible in hundreds of research laboratories or can be purchased for less than $200, and a standard cell phone camera can serve as a recording device. While we do not attempt a comprehensive survey of all possible mask designs, important general characteristics of face masks can be extracted. The experimental setup is simple and can easily be built and operated by non-experts. The described method allows for a relative comparison between different face masks and their transmission of droplets.

**Fig. 1.**
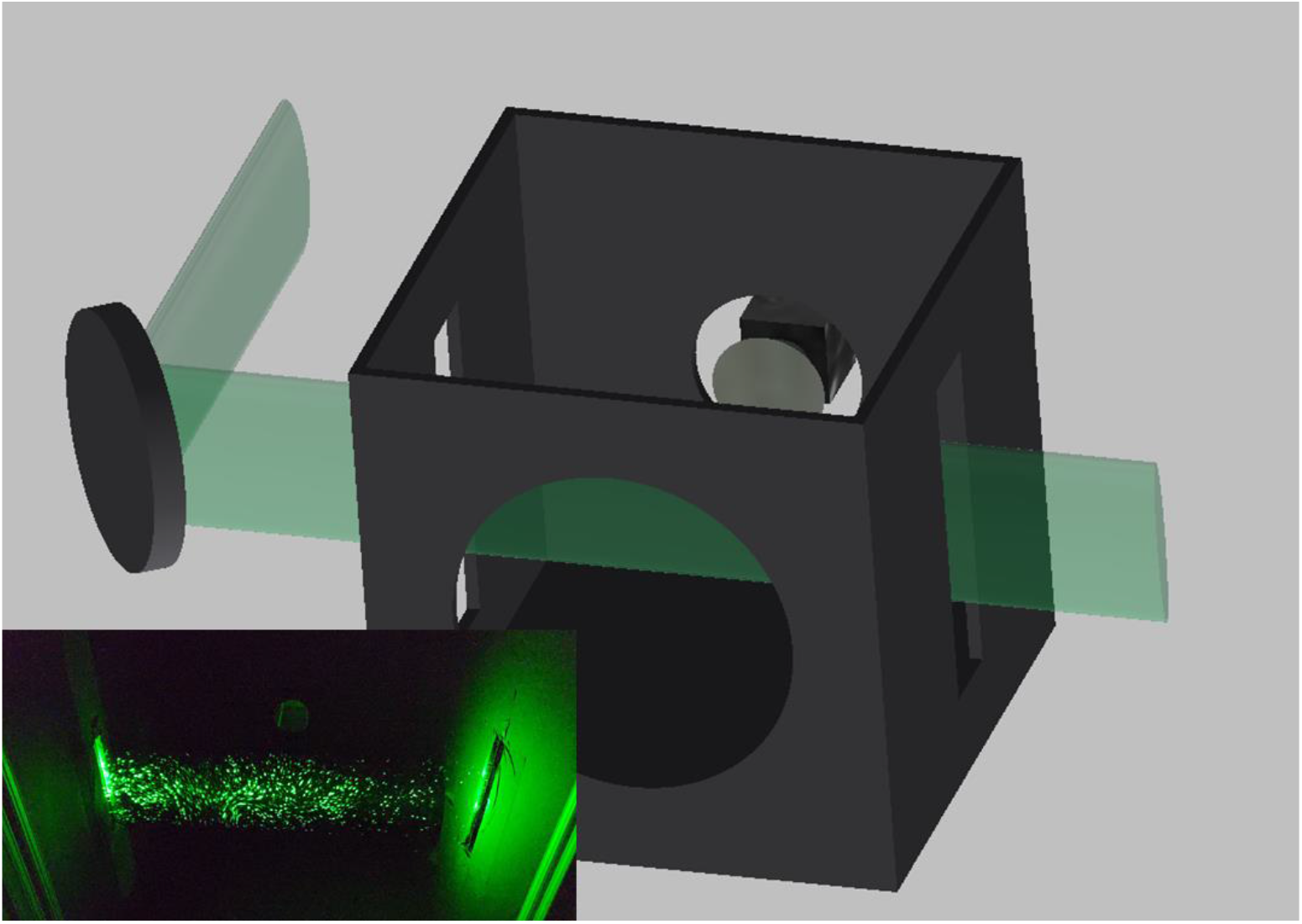
Schematic of the experimental setup. A laser beam is expanded vertically by a cylindrical lens and shined through slits in the enclosure. The camera is located at the back of the box, a hole for the speaker in the front. The inset shows scattering for water particles from a spray bottle with the front of the box removed.

## Results

We tested 12 commonly available masks or masks alternatives and one patch of mask material (see **Fig. 2** and **Table 1** for details). In addition, we recorded a control trial where the speaker wore no protective mask or covering. Each test was performed with the same protocol. The camera was used to record a video of approximately 40 seconds length to record droplets emitted while speaking. The first 10 s of the video serve as baseline. In the next 10 s, the mask wearer repeated the sentence “Stay healthy, people” five times (speech), after which the camera kept recording for an additional 20 s (observation). For each mask and for the control trial, this protocol was repeated 10 times. We use a computer algorithm (see Materials and Methods) to count the number of particles within each video.

**Table 1.** Face masks under investigation. This table lists the investigated face masks, mask alternatives, and mask material (masks are depicted in **Fig. 1)**.

| Mask, Name | Description |
| --- | --- |
| 1, 'Surgical' | Surgical mask, 3-layer |
| 2, 'N95' | N95 mask with exhalation valve |
| 3, 'Knitted' | Knitted mask |
| 4, 'PolyProp' | 2-layer polypropylene apron mask |
| 5, 'Poly/Cotton' | Cotton-polypropylene-cotton mask |
| 6, 'MaxAT' | 1-layer Maxima AT mask |
| 7, 'Cotton2' | 2-layer cotton, pleated style mask |
| 8, 'Cotton4' | 2-layer cotton, Olson style mask |
| 9, 'Cotton3' | 2-layer cotton, pleated style mask |
| 10, 'Cotton1' | 1-layer cotton, pleated style mask |
| 11, 'Fleece' | Gaiter type neck fleece |
| 12, 'Bandana' | Double-layer bandana |
| 'Swath' | Swath of mask material, polypropylene |
| 'None' | Control experiment, no mask |

**Fig. 2.**
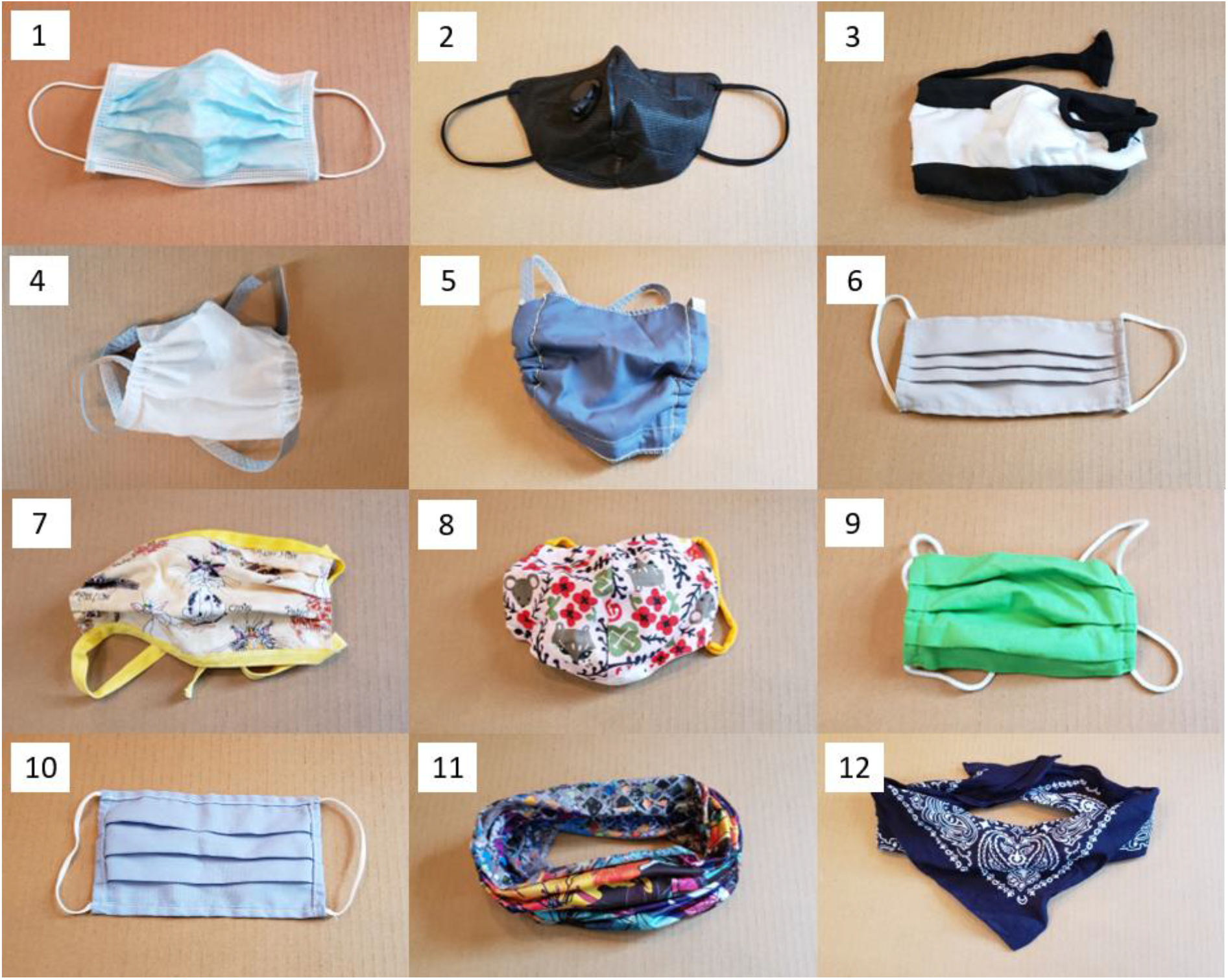
Pictures of face masks under investigation. We tested 12 different face masks or mask alternatives and one mask material (not shown).

The main result of our study is depicted in **Fig. 3 (A)**, where we show the total droplet count for each tested mask on a logarithmic scale. Data points and error bars represent the mean value and distribution standard deviation, respectively, of the total droplet count normalized to the control trial (no mask). For our control trial, the absolute droplet count was around 960. We measured a droplet transmission fraction ranging from 5% (surgical mask) to 110% (fleece mask, see discussion below) relative to the control trial. In **Fig. 3 (B)**, the time evolution of detected droplets is shown for three representative examples (one of the cotton masks, the bandana, and the control trial) – the data for all tested masks is shown in Supplementary Figure **S1**. The solid curves indicate the droplet transmission rate over time. For the control trial (green curve), the five distinct peaks correspond to the five repetitions of the operator speaking. In the case of speaking through a mask, there is a physical barrier, which results in a reduction of transmitted droplets and a significant delay between speaking and detecting particles. In effect, the mask acts as a temporal low pass filter, smoothens the droplet rate over time, and reduces the overall transmission. For the bandana (orange curve), the droplet rate is merely reduced by a factor of two and the repetitions of the speech are still noticeable. The effect of the cotton mask (blue curve) is much stronger. The speech pattern is no longer recognizable and most of the droplets, compared to the control trial, are suppressed. The shaded areas for all three curves display the cumulative particle count over time: the lower the curve, the more droplets are blocked by the mask.

**Fig. 3.**
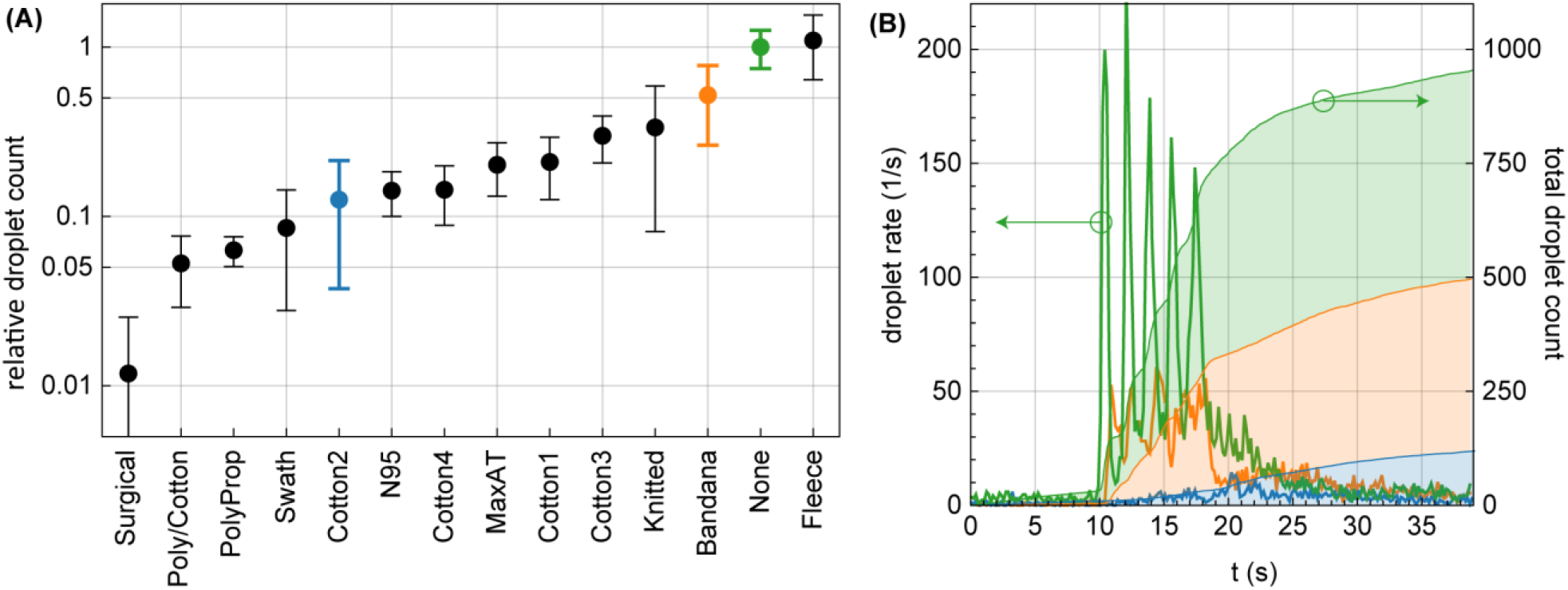
Droplet transmission through face masks. **(A)** Relative droplet transmission through the corresponding mask. Each data point represents the mean and standard deviation over 10 trials for the same mask, normalized to the control trial (no mask). **(B)** The time evolution of the droplet count (left axis) is shown for three representative examples, marked with the corresponding color in (A): No mask (green), Bandana (orange), and cotton mask (blue). The cumulative droplet count for these cases is also shown (right axis).

We notice that speaking through some masks (particularly the neck fleece) seemed to disperse the largest droplets into a multitude of smaller droplets (see supplementary figure **S2**), which explains the apparent increase in droplet count relative to no mask in that case. Considering that smaller particles are airborne longer than large droplets (larger droplets sink faster), the use of such a mask might be counterproductive. Furthermore, the performance of the N95 mask is likely affected by the exhalation valve, which opens for strong outwards airflow. While the valve does not compromise the protection of the wearer, it can decrease protection of persons surrounding the wearer.

## Discussion

The experimental setup is very straightforward to implement and the required hardware and software are ubiquitous or easily acquired. However, this simplicity does go along with some limitations that are discussed here.

First, our experimental implementation samples only a small part of the enclosure and hence some droplets that are transmitted through the masks might not be registered in the laser beam. This means that the droplet count reflect only a portion of all droplets, but as we perform the experiment with same initial conditions for all masks, the relative performance of the masks can be compared.

Second, the use of a cell phone camera poses certain limitations on detection sensitivity, i.e. the smallest recognizable droplet size. To estimate the sensitivity, we consider the light that is scattered by droplets passing through the laser beam. The amount of light scattered into the camera direction depends on the wavelength of light, the refractive index of the droplet, and its size (and shape). To estimate the light scattering of droplets into the camera as a function of their diameter we used the Python package PyMieScatt [12], which is an implementation of Lorenz-Mie theory (see [13] for a review). The result is visualized in **Fig. 4**. Panel (A) shows an example of the scattering distribution for 532 nm light scattered from a droplet of 5 μm diameter and a refractive index of water (n=1.33). In this example, the particle size is substantially bigger than the wavelength of the light (the so-called Mie regime). Almost all the light is scattered into the forward direction (0°) and very little into the direction of the camera (indicated by the shaded green cone around 90°). For the given camera acceptance angle, we display in Fig. 4 (B) the estimated number of photons per frame scattered into the cell phone camera aperture as a function of particle diameter. By illuminating the camera directly with an attenuated laser beam of known power, we determine the detection sensitivity. A minimum of about 75 photons (on a single camera pixel) or about 960 photons (spread over several pixels) per frame were required for the camera to detect a droplet (for details on the detection characterization, see the supplementary materials). Both detection thresholds are indicated by horizontal black lines in and the red shaded area in Fig. 4 (B). The more conservative detection threshold corresponds to a minimum detectable droplet size of 0.5 µm. The main limitation is the low collection efficiency of our small camera aperture - we currently capture only 0.01% of the full solid angle. An increased collection efficiency is possible with a larger relay lens in front of the camera, but this would come at the cost of a reduced field of view.

**Fig. 4.**
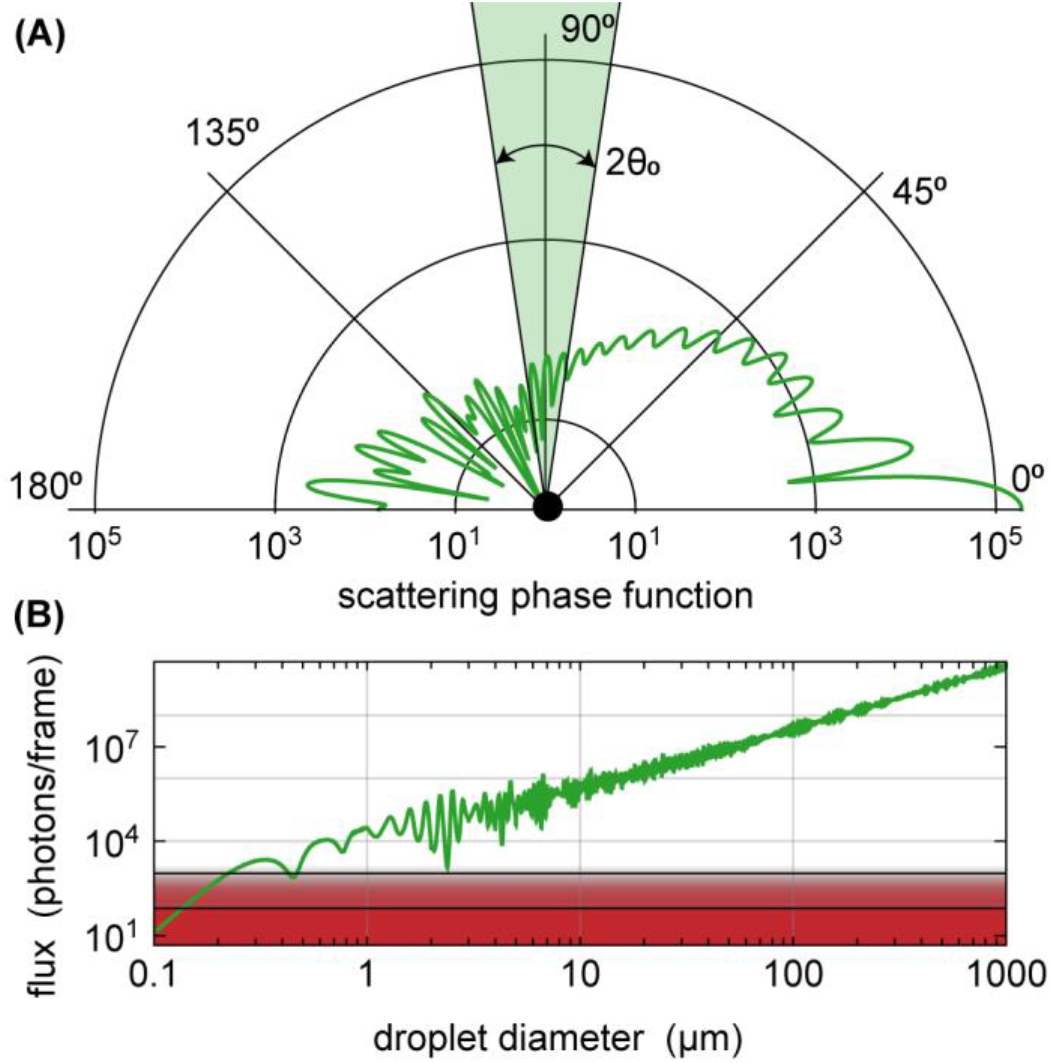
Light scattering properties. **(A)** Angle distribution (scattering phase function) for light scattered by a water droplet of 5 µm diameter for illumination with green laser light. Note the logarithmic radial scale. 0° is the forward direction, 180° the backward direction. The camera records at around 90°, indicated by the green segment (not to scale). (**B**) Calculated number of photons recorded by the camera in one frame as a function of the droplet diameter. The red shaded area and the two solid lines indicate the detection thresholds of the camera. For ideal conditions (all photons impinge on a single pixel), the camera requires at least about 75 photons per frame corresponding to a droplet diameter of 0.1 µm; for photons distributed over multiple pixels, the threshold is around 960 photons and correspond to a diameter of 0.5 µm.

Third, the use of a single cell phone camera also limits the achievable size resolution (currently 120 µm/pixel), given the large field of view that is required to image as many droplets as possible. This makes it unfeasible to directly measure the size of small (aerosol) droplets in our setup. However, while we cannot measure the size of droplets at or below the pixel resolution, we can still detect and count the smaller droplets, down to the sensitivity limit described above. For very large particles, the limited dynamic range of the camera also poses a challenge for determining the size, since pixels easily saturate and hence distort the shape of the recorded droplet. We want to point out that neither the limited pixel resolution nor the saturation affect the particle counts presented in Fig. 3. Choosing a higher quality camera and a smaller field of view, combined with a funnel setup to guide droplets towards the imaging area, would reduce the minimum observable size; so would approaches which use camera arrays to improve resolution without sacrificing sensitivity or field of view [14]. Keeping in mind these sizing limitations, we can still estimate the size distribution for the larger droplets (see supplementary figure S2 for a qualitative size plot), which presents some interesting observations such as the fleece performance mentioned earlier.

We should point out that our experiments differ in several ways from the traditional methods for mask validation, such as filtration efficiency of latex particles. As is apparent from the neck fleece study, liquid filtration (and subsequent particle size reduction) are more relevant than solid filtration. In addition, our method could inform attempts to improve training on proper mask use and help validate approaches to make existing masks reusable.

In summary, our measurements provide a quick and cost-effective way to estimate the efficacy of masks for retaining droplets emitted during speech for droplet sizes larger than 0.5 µm. Our proof-of-principle experiments only involved one speaker, but our setup allows for easy future checks of mask performance under a variety of conditions that affect the droplet emission rate, like different speakers, volume of speech [15], speech patterns [16], and other effects. This method can also test masks under other conditions, like coughing or sneezing. Improvements to the setup can increase sensitivity, yet testing efficiency during regular breathing likely will require complementing measurements with a conventional particle sizer. A further area of interest is the comparison of mask performance between solid particles and droplets, motivated by the observed liquid droplet breakup in the neck fleece and mask saturation by droplets, necessitating exchange in regular clinical practice.

## Materials and Methods

The optical setup we employed was recently used to demonstrate expulsion of liquid droplets during speech and for characterization of droplet residence times in air [1, 2]. A schematic of the setup is shown in **Fig. 1**. In short, a light sheet was shined through an enclosure where light scattering from particles traversing the light sheet was detected with the camera. To form the light sheet, a cylindrical lens transformed a green laser beam into an elliptical profile, which was directed through the enclosure. The laser source was a scientific pump laser (Millennia, Spectra-Physics; power 2 W, wavelength 532 nm), but suitable green lasers of similar powers are available for less than US $100; the scientific lasers have better specifications (higher beam pointing and intensity stability, better beam profile), but these advantages are irrelevant in this application. The light sheet at the center of the enclosure had a thickness of 4.4 mm and a vertical size of 78 mm (Gaussian 1/e^2^ intensity beam widths). The enclosure (L × W × H: 30 cm × 30 cm × 35 cm) was constructed out of (or lined with) black material to minimize stray light. The sides of the box had slits for entry and exit of the light sheet. The front of the box had an 18 cm diameter hole for the speaker – large enough for a person wearing a mask to speak into the box but small enough to prevent the face (or mask) from reaching the light sheet. In order to clear droplets from the box between experiments, laminar HEPA-filtered air was continuously fed into the box from above through a duct of cross section 25 cm × 25 cm. The supplied air was being expelled through the light sheet slits and the speaker hole. A slight positive pressure in the box cleared droplets and prevented dust from entering into the box from outside. On the back of the box, a cell phone (Samsung Galaxy S9) was mounted at a distance of 20 cm from the light sheet. Using the Android app “Open Camera” the frame size was set to 1920 × 1080 pixels, the focal distance to 20 cm, the exposure time to 1/50 s, and the frame rate to 30/s. At this focal distance, each camera pixel recorded an area of 120 µm × 120 µm at the position of the light sheet.

For each trial, the camera recorded scattered light from particles in the laser beam before the speech (∼10 s), during speech (∼10 s), and for a period of droplet clearing (∼20 s). The speech consisted of five repetitions of the phrase “Stay healthy, people,” spoken by a male test person with a strong voice but without shouting. Each trial was repeated ten times and the speaker drank a sip of water in between to avoid dehydration. Furthermore, for the masks that showed substantial amounts of detected particles (knitted, cotton, fleece, and bandana), we conducted additional tests by repeatedly puffing air from a bulb through the masks, rather than speech from an experimenter. These control trials with air puffs confirmed that we recorded droplets emitted by the speaker, not dust from the masks.

The goal of the analysis is to compare the efficacy of different masks by estimating the total transmitted droplet count. Towards this end, we need to identify droplets in the video and discriminate between droplets and background or noise. For convenience, analysis of the videos was performed with “Mathematica” (Wolfram Research), but use of a commercial package does not pose any general restriction since almost every high-level programming language (e.g. Python) offers the same functionality. From all videos, we removed a weak background that originated from the light sheet itself and from stray light and diffuse reflections from the experimenter’s face. We then binarized all frames with a common threshold that discriminates between scattered light from droplets and background signal and/or noise. Then, a feature detection algorithm is applied to each frame, which returns the center of mass positions, and major axis and minor axis length of the best-fit ellipse for every droplet. Note that the major and minor axis returned by the algorithm are not a direct measure of the droplet size, but a measurement of the amount of light scattered by the particle into the camera aperture (binary diameter). Furthermore, the major axis length is increased due to particle motion during the camera exposure time. Due to the small dynamic range of the camera (8-bit), most droplets saturate the camera. However, the axes lengths returned by the algorithm can still be used for a qualitative droplet size estimation: a bigger droplet scatters more light than a smaller droplet. This insight is important to interpret the result of the neck fleece. The neck fleece has a larger transmission (110%, see Fig. 3 (A)) than the control trial. We attribute this increase to the neck fleece dispersing larger droplets into several smaller droplets, therefore increasing the droplet count. The histogram of the binary diameter for the neck fleece supports this theory (see Fig. S2).

If a droplet passes through the light sheet in a time shorter than the inverse frame rate, it will appear only in a single video frame. However, if the droplet spends more time in the light sheet, the droplet will appear in multiple frames. To avoid double-counting droplets in consecutive frames, we use a basic algorithm to distinguish between single-frame particles and multi-frame trajectories. The algorithm compares the distance between droplets in consecutive frames and assigns two droplets to a trajectory if their distance is smaller than a threshold value, or counts them as individual droplets if their distance is larger than the threshold. The threshold value was empirically chosen to be 40 pixels. An example result of the algorithm is shown in supplementary figure **S3**, which shows a projection of 10 consecutive frames. Every droplet recognized by the algorithm is highlighted by an ellipsoid, labeled with the frame number. Droplets that belong to the same trajectory are highlighted in the same color.

## Data Availability

All movie files are available freely at the Duke Research Data Repository at http://doi.org/10.7924/r4kp81n9j.

## Supplementary Materials

Materials and Methods

Fig. S1. Time evolution of droplet rate and total droplet count.

Fig. S2. Qualitative size histogram.

Fig. S3. Example frame projection demonstrating particle trajectories.

Fig. S4. Camera calibration and sensitivity.

## Acknowledgments

We thank Mathias Fischer for providing the sketch in Fig. 1, and Shannon Eriksson and Jake Lindale for valuable discussions.

## Author contributions

M.C.F. and E.P.F. performed the experiments, D.G. performed the data analysis, I.H. and E.W. procured the masks, and W.S.W. provided expertise. M.C.F supervised the project. All authors were involved in data interpretation and manuscript preparation.

## References

1. Stadnytskyi, V., et al., The airborne lifetime of small speech droplets and their potential importance in SARS-CoV-2 transmission. Proceedings of the National Academy of Sciences, 2020. 117(22): p. 11875–11877.

2. Anfinrud, P., et al., Visualizing Speech-Generated Oral Fluid Droplets with Laser Light Scattering. New England Journal of Medicine, 2020. 382(21): p. 2061–2063.

3. Belkin, N.L., The Evolution of the Surgical Mask: Filtering Efficiency versus Effectiveness. Infection Control and Hospital Epidemiology, 1997. 18(1): p. 49–57.

4. Dharmadhikari, A.S., et al., Surgical face masks worn by patients with multidrug-resistant tuberculosis: impact on infectivity of air on a hospital ward. Am J Respir Crit Care Med, 2012. 185(10): p. 1104–9.

5. WHO, Advice on the use of masks in the context of COVID-19. 2020, World Health Organization.

6. Klompas, M., et al., Universal Masking in Hospitals in the Covid-19 Era. New England Journal of Medicine, 2020. 382(21): p. e63.

7. Chu, D.K., et al., Physical distancing, face masks, and eye protection to prevent person-to-person transmission of SARS-CoV-2 and COVID-19: a systematic review and meta-analysis. The Lancet, 2020.

8. Leung, N.H.L., et al., Respiratory virus shedding in exhaled breath and efficacy of face masks. Nature Medicine, 2020. 26(5): p. 676–680.

9. Bunyan, D., et al., Respiratory and facial protection: a critical review of recent literature. Journal of Hospital Infection, 2013. 85(3): p. 165–169.

10. Ong, J.J.Y., et al., Headaches Associated With Personal Protective Equipment – A Cross-Sectional Study Among Frontline Healthcare Workers During COVID-19. Headache: The Journal of Head and Face Pain, 2020. 60(5): p. 864–877.

11. Konda, A., et al., Aerosol Filtration Efficiency of Common Fabrics Used in Respiratory Cloth Masks. ACS Nano, 2020. 14(5): p. 6339–6347.

12. Sumlin, B.J., W.R. Heinson, and R.K. Chakrabarty, Retrieving the aerosol complex refractive index using PyMieScatt: A Mie computational package with visualization capabilities. Journal of Quantitative Spectroscopy and Radiative Transfer, 2018. 205: p. 127–134.

13. Gouesbet, G. and G. Gréhan, Generalized Lorenz-Mie Theories. 2nd ed. 2017: Springer International Publishing.

14. Zhong, L., et al. Depth tracking using a multi-aperture microscope. in Imaging and Applied Optics. 2019. Optical Society of America.

15. Asadi, S., et al., Aerosol emission and superemission during human speech increase with voice loudness. Scientific Reports, 2019. 9(1): p. 2348.

16. Asadi, S., et al., Effect of voicing and articulation manner on aerosol particle emission during human speech. PLOS ONE, 2020. 15(1): p. e0227699.

